# Exploring the niche of *Rickettsia montanensis* (Rickettsiales: Rickettsiaceae) infection of the American dog tick (Acari: Ixodidae), using multiple species distribution model approaches

**DOI:** 10.1101/2020.09.29.20204149

**Authors:** Catherine A. Lippi, Holly D. Gaff, Alexis L. White, Heidi K. St. John, Allen L. Richards, Sadie J. Ryan

## Abstract

The American dog tick, *Dermacentor variabilis* (Say), is a vector for several human disease causing pathogens such as tularemia, Rocky Mountain spotted fever, and the understudied spotted fever group rickettsiae (SFGR) infection caused by *Rickettsia montanensis*. It is important for public health planning and intervention to understand the distribution of this tick and pathogen encounter risk. Risk is often described in terms of vector distribution, but greatest risk may be concentrated where more vectors are positive for a given pathogen. When assessing species distributions, the choice of modeling framework and spatial layers used to make predictions are important. We first updated the modeled distribution of *D. variabilis* and *R. montanensis* using MaxEnt, refining bioclimatic data inputs, and including soils variables. We then compared geospatial predictions from five species distribution modeling (SDM) frameworks. In contrast to previous work, we additionally assessed whether the *R. montanensis* positive *D. variabilis* distribution is nested within a larger overall *D. variabilis* distribution, representing a fitness cost hypothesis. We found that 1) adding soils layers improved the accuracy of the MaxEnt model; 2) the predicted ‘infected niche’ was smaller than the overall predicted niche across all models; and 3) each model predicted different sizes of suitable niche, at different levels of probability. Importantly, the models were not directly comparable in output style, which could create confusion in interpretation when developing planning tools. The random forest (RF) model had the best measured validity and fit, suggesting it may be most appropriate to these data.

## Introduction

The American dog tick (*Dermacentor variabilis*) is primarily distributed east of the Rocky Mountains and the Pacific coastal region in the USA, where it is a known vector of the pathogens that cause both tularemia, (*Francisella tularensis*) and Rocky Mountain spotted fever (RMSF) (*Rickettsia rickettsii*). Both diseases can be fatal if left untreated, and therefore understanding the risk of exposure to *D. variabilis* bites is an essential part of public health planning. In addition to these more well-known vector-borne diseases, *D. variabilis* can also transmit *Rickettsia montanensis*, a spotted fever group rickettsiae (SFGR). *Rickettsia montanensis* was previously thought to be nonpathogenic in humans (Baldridge et al. 2010) but has more recently been implicated as the agent in an afebrile rash illness (McQuiston et al. 2012). In addition to the potential for human pathogenicity, *R. montanensis* may play an interesting role in the manifestation of other SFGR dynamics by inhibiting tick coinfection of another *Rickettsia* spp., or conferring antigenic responses in humans exposed to *R. montanensis*, providing immunity (partial or complete) to other SFGR pathogens (Baldridge et al. 2010). This has potential implications for the transmission cycles of other SFGR, namely *R. rickettsii*, which occurs sympatrically with *R. montanensis. Rickettsia montanensis* infections may also affect SFGR disease surveillance and case detection. In 2010, the Centers for Disease Control (CDC) designated a new category for reporting rickettsial diseases to reflect diagnostic uncertainty in cases (CDC 2010, 2019a). The new reporting group, Spotted Fever Rickettsiosis (SFR), includes cases of RMSF, Pacific Coast tick fever, *Rickettsia parkeri* rickettsiosis (Tidewater spotted fever), and rickettsialpox. The number of SFR cases reported in the USA has shown a generally increasing trend since 2010, with more than 6,200 cases reported in 2017 (CDC 2019a). It is plausible that *R. montanensis* is the agent responsible for some of these SFR cases, as commonly used serologic tests are not able to differentiate rickettsial pathogens because of immunological cross-reactivity (CDC 2019a, Nicholson and Paddock 2019). The public health implications of *R. montanensis* infections, both for human health and case surveillance, has fueled interest in investigating this pathogen.

The previous assumption of *R. montanensis* being nonpathogenic in humans, and non-specific point of care tests for SFGR has led to few publications on this pathogen and its known or potential distribution, as pointed out by Hardstone, Yoshimizu and Billeter (2018). Estimation of the geographic distribution for pathogens and their vectors is a crucial component in the development of public health agency policies and recommendations. In 2016, St. John and colleagues (St. John et al. 2016) conducted a study to describe the predicted distributions of *R. montanensis* positive and negative *D. variabilis* in the United States using the MaxEnt modeling environment to generate species distribution models (SDMs). Species distribution models have become increasingly prevalent in the disease ecology literature, with examples spanning a range of infectious disease systems, spatial scales, and geographic foci (Gurgel-Gonçalves et al. 2012, Blackburn et al. 2017, Lippi et al. 2019). This methodology has been embraced as an accessible means of quickly estimating the geographic range of a given pathogen or vector, which is used in many instances to infer risk of exposure.

Species distribution models are attractive from a logistical standpoint for mapping potential exposure to vectors as they provide a means of estimating suitable geographic ranges with presence-only data, which are often available through public surveillance networks. Briefly, SDMs are made by correlating location records of species occurrence with underlying environmental conditions in a geospatial modeling environment, and the model is then projected to unsampled portions of the landscape (Peterson and Soberón 2012). There is now a range of modeling algorithms and freely available software packages that, when coupled with the availability of georeferenced occurrence records, have made SDMs commonly used across disciplines in recent years (Townsend Peterson et al. 2007, Elith et al. 2008, Phillips and Dudík 2008, Elith and Leathwick 2009, Evans et al. 2011, Naimi and Araújo 2016). Nevertheless, caution must be exercised when modeled distributions are put into a health advisory context. Consensus in SDMs is notoriously difficult to achieve, as discrepancies in potential distributions may arise from choice of modeling method, user-specified parameters, selection of environmental predictors, and biases in data inputs (Carlson et al. 2018).

The objectives of this study were as follows: i) expand the methodology used to generate the predicted range map of *D. variabilis* presented in St. John et al. (St. John et al. 2016) by creating SDMs in MaxEnt with an updated and refined set of environmental predictors; ii) explore differences across four additional SDM algorithms; and iii) compare potential geographic distributions of *D. variabilis* and the subset of *D. variabilis* that tested positive for *R. montanensis* to assess if there were any appreciable differences in the ‘infected’ niche.

## Methods

### Presence data

Locations of *D. variabilis* in the US from 2002-2012, which tested both positive and negative for *R. montanensis*, are described in St John et al. (2016). Data were openly available through VectorMap (http://vectormap.si.edu/dataportal/), a project of the Walter Reed Bioinformatics Unit, housed at the Smithsonian Institution Washington DC (St. John et al. 2016). These data were collected primarily through reports to United States military installations and as part of passive vector surveillance studies. Prior to implementing modeling procedures, we conducted data thinning on species occurrence points via the spThin package in R (ver. 3.6.1) (R Core Team 2019), which uses a spatial thinning algorithm to randomize the removal of occurrence locations within a specified distance threshold (Aiello-Lammens et al. 2015). The resulting dataset retained spatially unique records of species presence within 10km, the spatial resolution of the study. Locations of tick occurrences in these data were provided as either the location of collection or the associated medical treatment facility. Correlative SDMs are susceptible to the effects of geographic sampling biases, where overrepresented locations may erroneously drive associations between environmental conditions at oversampled locations and species occurrence (Aiello-Lammens et al. 2015). Spatial thinning of occurrences at the chosen spatial resolution minimizes the potential effects of sampling bias in this dataset, where locations near reporting medical facilities may be overrepresented.

#### Environmental data layers

Species distribution models, of the type we present in this study, require gridded environmental data layers as input for building models and making spatial predictions. For comparability with the previous study (St. John et al. 2016), and to maintain consistency across algorithms in this paper, we used interpolated bioclimatic (BIOCLIM) layers from WorldClim.org at a 10km resolution selected to match the spatial resolution of tick occurrence data (Fick and Hijmans 2017). The 19 BIOCLIM variables consist of long-term averages of temperature, precipitation, and associated measures of extremes and seasonality.

In addition to the original layer set from the previous study, we chose to add soils layers to our candidate environmental variables, as ticks are frequently found in the leaf-litter or debris and considered largely soil-dwelling organisms (Burtis et al. 2019). The International Soil Reference Information Centre (ISRIC) SoilGrids product provides a global suite of 195 standard numeric and taxonomic soil descriptors at seven standard depths (Hengl et al. 2017). As ticks are sensitive to abiotic soil attributes such as soil moisture (Burtis et al. 2019), we selected two layers of soil data for inclusion in the candidate variable set for model building; soil organic carbon density and available soil water capacity until wilting point to describe the potential water capacity and retention of soils. A standard depth of 0cm was chosen to approximate the surface and leaf litter conditions that ticks may encounter in the environment. Gridded soil layer products, aggregated to a spatial resolution of 10km with the GDAL software package, were used to match the resolution of the study (GDAL/OGR contributors 2020).

Collinearity in environmental predictor variables is a well-described issue affecting SDM output, potentially increasing model instability and uncertainty in predictions (De Marco and Nóbrega 2018). We reduced collinearity in environmental variable inputs via variance inflation factor (VIF), wherein only those layers with values below a specified threshold (th=10) were used in model building (Chatterjee and Hadi 2006).

##### Model implementation

We used the ‘sdm’ package in R (ver. 3.6.2) to fit and spatially project SDMs for positive ticks and the combined dataset of positive and negative ticks (Naimi and Araújo 2016). The sdm package provides a flexible modeling platform for building SDMs, assessing model accuracy, and projecting output. Choice of modeling method can result in drastically different predictions of species ranges. To assess variation in potential distributions as an artifact of methodology, we used five commonly implemented modeling algorithms for estimating species ranges. These included two regression methods, generalized linear model (GLM) and generalized additive model (GAM), and three machine learning methods: maximum entropy (MaxEnt), random forests (RF), and boosted regression trees (BRT) (McCullagh and Nelder 1998, Breiman 2001, Wood 2006, Elith et al. 2008, Phillips and Dudík 2008). We additionally created an SDM modeling ensemble, where predictions were weighted by accuracy metrics and averaged across methods (Araujo and New 2007). Model ensembles have been criticized due to performance issues and poor reporting practices (Hao et al. 2019) but are widely used. In spite of known issues, we included the ensemble method for the purposes of comparison, as SDM ensembles are prolific in the literature and provide a useful tool for combining the results from the various model approaches.

Model parameterization also heavily influences resulting predictions of species’ distributions. While model settings vary depending on method, they are generally chosen based on known attributes of the target species (e.g. physiological thermal limits, nonlinear responses to environmental drivers, etc.), intended application of results (e.g. hypothesis testing, biological interpretation of niche, designing interventions, etc.), or to address issues with bias and small sample size (Merow et al. 2013, Morales et al. 2017). *Dermacentor variabilis* are habitat generalists, and in contrast with many arthropod systems, little is known for acarids particularly regarding quantitative relationships with environmental conditions. We therefore used default modeling parameters for each method as defined in the sdm modeling platform. Five hundred model replications were run for each method, using a random subsampling of occurrence records (80%) for each model. Because we used presence-only data of species occurrences, pseudo-absences (n=1,000) were randomly generated throughout the study region within the sdm modeling procedure for each model run. Accuracy metrics were derived via a random subsampling (20%) of testing data, withheld from the model building process. Four measures of model accuracy were used to assess model output. These included the receiver operator characteristic (ROC) curve with area under the curve (AUC), true skill statistic (TSS), model deviance, and mean omission (i.e. false negatives).

Spatial predictions were made by projecting the mean of all 500 models for a given method onto the study area. Overlap in geographic predictions between ticks positive for *R. montanensis* and the full dataset of tick occurrences (i.e. *R. montanensis* positive and negative ticks) was assessed by reclassifying modeled probabilities as binary geographic distributions (i.e. presence and absence) in ArcMap (ver.10.4), where raster cells with predicted occurrence ≥ 50% were considered present (ESRI 2016). Reclassified distributions were combined using the ‘Raster Calculator’ tool in the Spatial Analyst extension of the program ArcMap, allowing for the visualization of range overlap between datasets.

## Results

The full dataset of georeferenced *D. variabilis* occurrences downloaded from VectorMap was comprised of 3,771 records, 135 of which had tested positive for *R. montanensis*. Spatial thinning of occurrence records resulted in 432 unique locations of *D. variabilis*, where a subset of 44 records was positive for *R. montanensis* (Fig. 1). Models for both the full set of *D. variabilis* occurrence records, and the pathogen positive subset, were built with a reduced set of VIF-selected environmental variables which included annual mean temperature, mean diurnal temperature range, temperature seasonality, mean temperature of the wettest quarter, mean temperature of the driest quarter, precipitation seasonality, precipitation of the warmest quarter, precipitation of the coldest quarter, soil organic carbon density, and available soil water capacity (Table 1).

**Table 1.** Environmental input datasets used in model building selected via variable inflation factor (VIF).

| Environmental Variable (unit) | Coded Variable Name | Data Source |
| --- | --- | --- |
| Annual Mean Temperature (°C) | Bio 1 | Bioclim |
| Mean Diurnal Range (°C) | Bio 2 | Bioclim |
| Temperature Seasonality | Bio 4 | Bioclim |
| Mean Temp of Wettest Quarter (°C) | Bio 8 | Bioclim |
| Mean Temp of Driest Quarter (°C) | Bio 9 | Bioclim |
| Precipitation Seasonality | Bio 15 | Bioclim |
| Precip of Warmest Quarter (mm) | Bio 18 | Bioclim |
| Precip of Coldest Quarter (mm) | Bio 19 | Bioclim |
| Soil Organic Carbon Density | OC Dens | ISRIC |
| Available Soil Water Capacity Until Wilting | WWP | ISRIC |

**Fig. 1.**
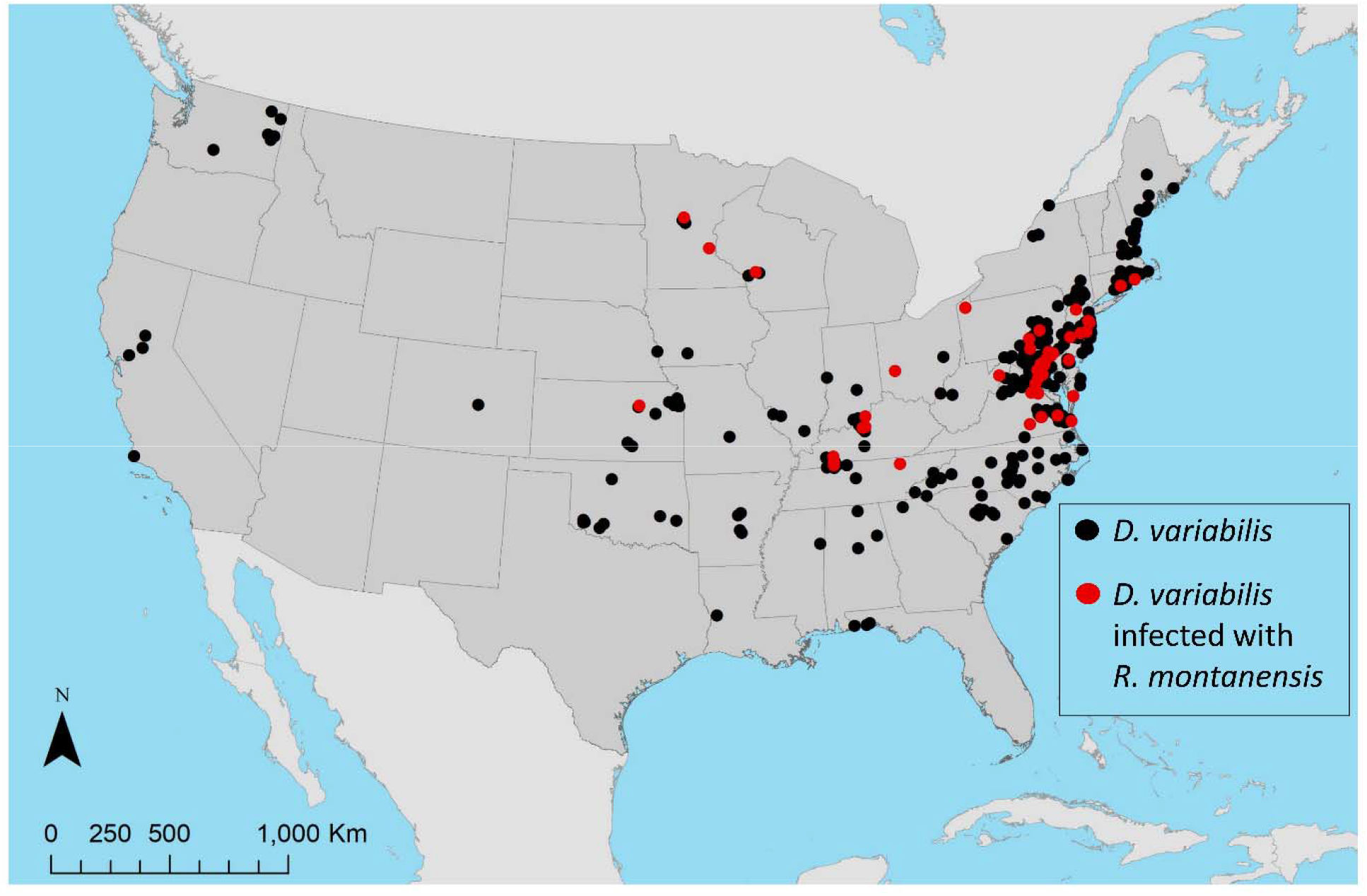
Occurrence records for *D. variabilis*, and *D. variabilis* infected with *R. montanensis*, used in building species distribution models (SDMs).

Accuracy metrics for averaged SDMs produced with each modeling method are presented in Table 2. The averaged model built with RF had the highest predictive power, relative to low deviation and omission error, for the full dataset (AUC=0.96, TSS=0.79, deviance=0.47, mean omission=0.11) and subset of positive ticks (AUC=0.93, TSS=0.81, deviance=0.20, mean omission=0.10). MaxEnt models also performed well, albeit with higher mean omission error for the full dataset (AUC=0.95, TSS=0.83, deviance=0.23, mean omission=0.12) and subset of positive ticks (AUC=0.95, TSS=0.77, deviance=0.66, mean omission=0.12). Averaged models produced with GLM for full dataset (AUC=0.90, TSS=0.70, deviance=0.80, mean omission=0.15) and positive subset (AUC=0.92, TSS=0.76, deviance=0.23, mean omission=0.15), and BRT for the full dataset (AUC=0.90, TSS=0.69, deviance=0.89, mean omission=0.15) and positive subset (AUC=0.91, TSS=0.77, deviance=0.27, mean omission=0.13) had relatively lower performance, with lower accuracy metrics and higher error compared to other methods.

**Table 2.** Accuracy metrics for species distribution models of *Dermacentor variabilis* ticks built with five modeling methods including generalized linear model (GLM), maximum entropy (MaxEnt), generalized additive model (GAM), random forests (RF), and boosted regression trees (BRT).

| Method | Dataset | AUC | TSS | Deviance | Mean Omission |
| --- | --- | --- | --- | --- | --- |
| GLM | Positive | 0.92 | 0.76 | 0.23 | 0.15 |
|  | All | 0.90 | 0.70 | 0.80 | 0.15 |
| GAM | Positive | 0.92 | 0.79 | 0.70 | 0.11 |
|  | All | 0.95 | 0.79 | 0.55 | 0.12 |
| MaxEnt | Positive | 0.95 | 0.83 | 0.23 | 0.12 |
|  | All | 0.95 | 0.77 | 0.66 | 0.12 |
| BRT | Positive | 0.91 | 0.77 | 0.27 | 0.13 |
|  | All | 0.90 | 0.69 | 0.89 | 0.15 |
| RF | Positive | 0.93 | 0.81 | 0.20 | 0.10 |
|  | All | 0.96 | 0.79 | 0.47 | 0.11 |

The overall pattern of estimated geographic distributions resulting from models built with all *D. variabilis* records was similar across methods, where the highest probabilities of suitable habitat were predicted in eastern United States, with some area in the West also identified as potentially suitable (Fig. 2). However, estimated probabilities of occurrence varied greatly across methods, with BRT (maximum 66.68%) and MaxEnt (maximum 67.08%) yielding generally low probabilities, and GLM (maximum 97.81%), GAM (maximum 99.60%), and RF (maximum 99.98%) producing models with generally higher probabilities of occurrence. The averaged ensemble of model predictions yielded intermediate probabilities (maximum 81.38%). *D. variabilis* was predicted across models to occur with relatively high probability in the northeastern United States, including areas in the states of Delaware, Pennsylvania, New Jersey, Connecticut, Maryland, Massachusetts, and Maine, and in the southern states of Virginia, West Virginia, Kentucky, Tennessee, North Carolina, and South Carolina. Lower probabilities of occurrence (i.e. < 50%), spanning much of the Midwest and limited areas in the West, were also consistent across models. The BRT model estimates the potential range of *D. variabilis* to extend across the entire North American continent, albeit with very low probability ranging 20-30%.

**Fig. 2.**
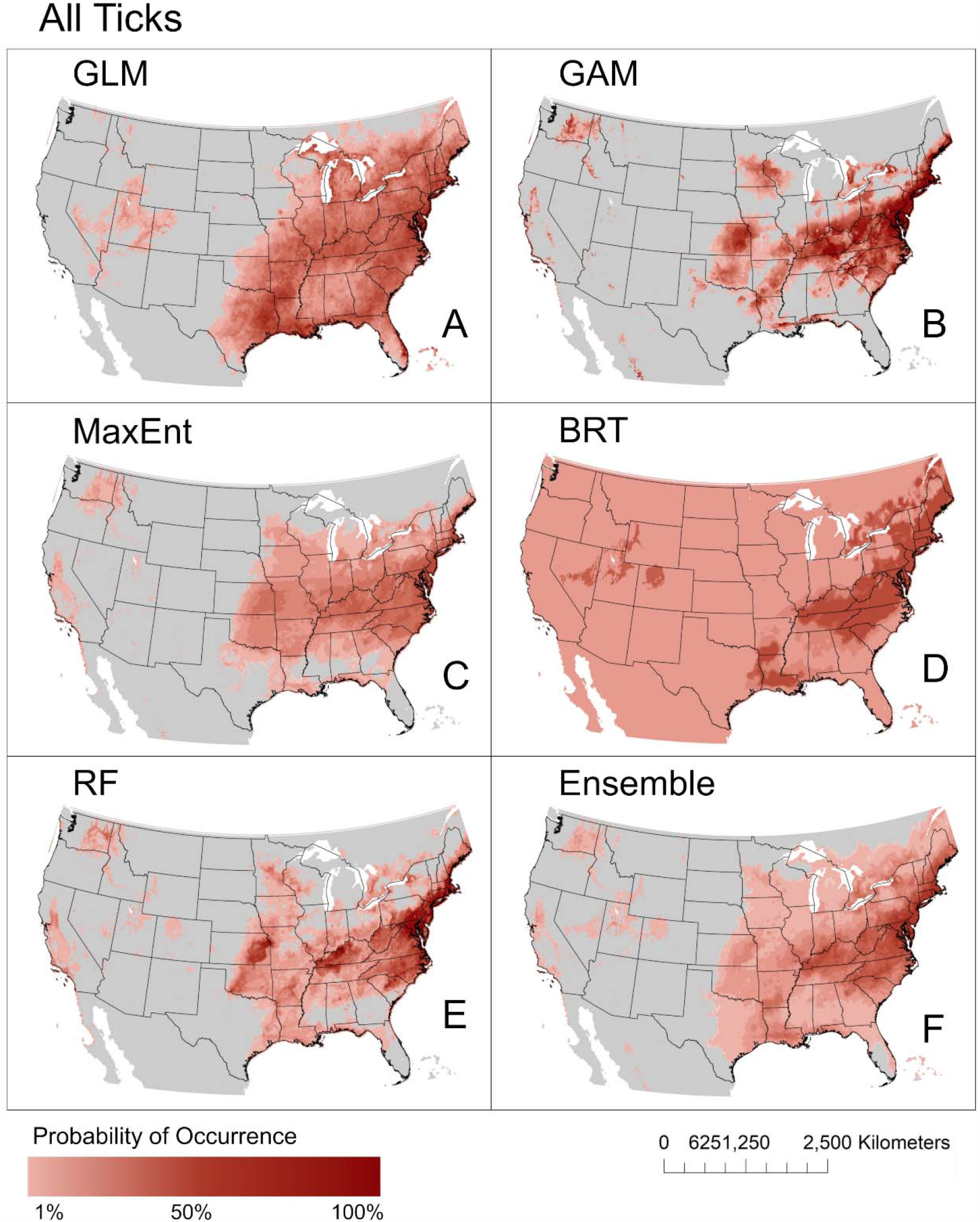
Predicted geographic distributions of *D. variabilis* ticks. Distributions were estimated using five common modeling methods including generalized linear model (GLM, A), generalized additive model (GAM, B), maximum entropy (MaxEnt, C), boosted regression trees (BRT, D), random forests (RF, E), and a weighted ensemble of these five methods (F).

The predicted distribution of *D. variabilis* which tested positive for *R. montanensis* was geographically constrained compared to the distributions estimated with the full dataset (Fig. 3).

**Fig. 3.**
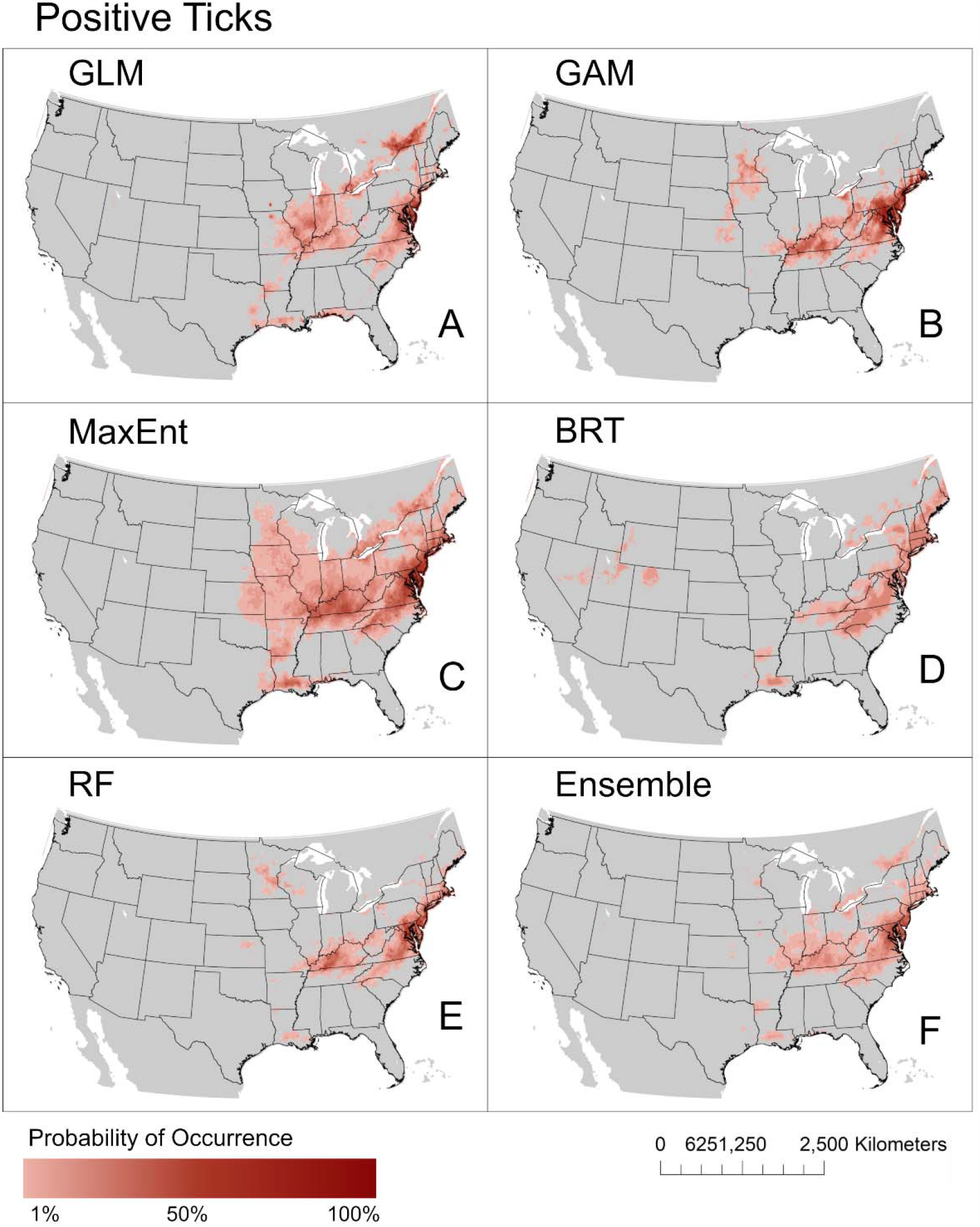
Predicted geographic distributions of *D. variabilis* ticks infected with *R. montanensis*. Distributions were estimated using five common modeling methods including generalized linear model (GLM, A), generalized additive model (GAM, B), maximum entropy (MaxEnt, C), boosted regression trees (BRT, D), random forests (RF, E), and a weighted ensemble of these five methods (F).

This pattern of restricted distribution was observed for all projected models, regardless of methodology. Four models (GLM, GAM, MaxEnt, and RF) project the highest probability for the occurrence of positive ticks in eastern states. The GAM, MaxEnt, and RF models predicted larger geographic extents for positive ticks, where high probabilities of occurrence (> 50%) spanned portions of states including Virginia, Maryland, Delaware, Pennsylvania, Connecticut, Rhode Island, and Massachusetts. The potential range of positive ticks in the East predicted via GLM were further restricted, where high probabilities for occurrence were observed in Virginia, Maryland, and New Jersey. Portions of Kentucky were also predicted to be suitable for positive ticks with high probability with three models (GAM, MaxEnt, and RF).The estimated range of positive ticks generated with BRT also included areas spanning states in the South, the Northeast, and some locations in the West. However, maximum probabilities of occurrence for positive ticks predicted with the BRT model are very low, not exceeding 40%.

Environmental factors that contributed the most to model averages for *D. variabilis* presence varied across methods. The top contributing environmental variable was precipitation seasonality for the GLM (81%), BRT (84%), and RF (26%) models; mean diurnal temperature range (30%) and mean annual temperature (25%) contributed the most to GAM models; mean annual temperature (28%) was most important for MaxEnt models (STable 1). Variable response curves varied greatly between models, but generally indicate that suitability is predicted when the precipitation in the warmest quarter is high and variation in precipitation seasonality is low (SFig. 1). Precipitation seasonality was the top contributing environmental factor for predictions of ticks testing positive for *R. montanensis* across all methods and was the single most important variable for MaxEnt (72%), BRT (98%), and RF (25%) models. Precipitation seasonality (99%), mean annual temperature (63%), temperature seasonality (60%), and precipitation of the warmest quarter (51%) were important variables in GLM models; precipitation seasonality (68%) and precipitation of the warmest quarter (50%) were important variables in GAM models (STable 2). Low variation in precipitation seasonality was the only common variable response across models of ticks positive for *R. montanensis* (SFig. 2).

Applying a conservative threshold for presence (i.e. where probabilities of occurrence less than 50% were considered unsuitable habitat) highlights where potential *D. variabilis* habitat occurs with the greatest probability (Fig. 4). High model predictions for presence of *D. variabilis* are predominantly concentrated in the eastern US across methods. Ticks positive for *R. montanensis* were predicted in geographically limited areas within the broader range of *D. variabilis* in all projected models, with the exception of the BRT model, which did not produce any probabilities for positive tick occurrences that met the threshold for presence (Fig. 4D). Restricted geographic distribution of positive ticks within the broader range of *D. variabilis* was also predicted in the model ensemble, although these were distinct potential ranges that did not overlap in areas with high probabilities (Fig. 4F).

**Fig. 4.**
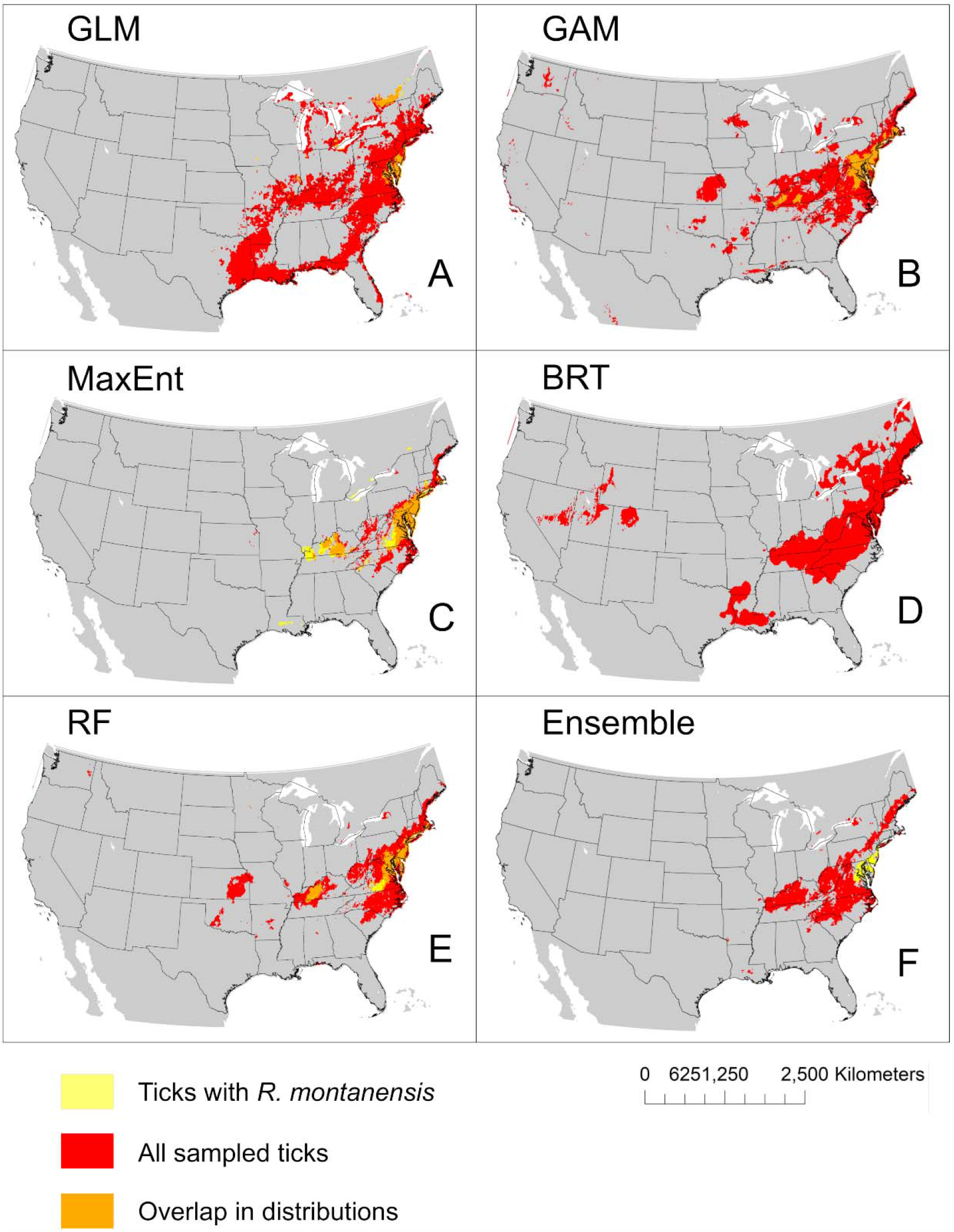
Overlap in the predicted geographic distributions (probability of occurrence > 50%) for *D. variabilis* ticks and *D. variabilis* ticks positive for *R. montanensis* infections. Distributions were estimated using five common modeling methods including generalized linear model (GLM, A), generalized additive model (GAM, B), maximum entropy (MaxEnt, C), boosted regression trees (BRT, D), and random forests (RF, E), and a weighted ensemble of these five methods (F).

## Discussion

The potential geographic ranges of *D. variabilis* produced in this study are largely in concurrence with maps of potential tick exposure issued by health authorities, where the ticks are reported to be widely distributed east of the Rocky Mountains, with limited distribution along the Pacific Coast (CDC 2019b). However, every modeling method, except for BRT, yielded geographic predictions for occurrence that were limited in area compared to general range maps for *D. variabilis*. These restrictions are more pronounced when locations with low probabilities of occurrence are omitted from mapped results, excluding known areas of occurrence in southern, midwestern, and Pacific coastal states (Fig. 4). *Dermacentor variabilis* with confirmed *R. montanensis* infections had constrained potential ranges within the broader geographic extent of *D. variabilis*. The potential range of positive *D. variabilis* modeled with a reduced variable set via MaxEnt in this study mirrors the results presented in St. John et al. (2016), where the highest probabilities of occurrence for positive ticks were focused in the Northeast and the Midwest, within the predicted range of *D. variabilis* that tested negative for *R. montanensis*. This restricted spatial pattern of predicted positive tick occurrences was also observed to varying degrees in models produced with three other methods (GLM, GAM, and RF).

*Dermacentor variabilis* is a habitat generalist, capable of exploiting a wide range of environmental conditions and host species (Sonenshine 1993). The bioclimatic variables that drove suitability predictions varied greatly between models, possibly reflecting the generalist life history of *D. variabilis*, and its ability to withstand conditions that would significantly limit other arthropod vectors. Nevertheless, indicators of seasonality, both in temperature and precipitation, were typically represented in averaged models, possibly reflecting the phenology of tick reproductive cycles. The life history of this vector may also elucidate the observed discrepancies in overall predicted range for *D. variabilis* presence versus the subset of pathogen positive tick occurrences. The restricted range of ticks positive for *R. montanensis*, within the larger area of suitable *D. variabilis* habitat is possibly indicative of an underlying range restriction in host availability. The hosts involved in zoonotic transmission cycles for *R. montanensis* are unknown, as they are with many tick-borne rickettsiae (Parola et al. 2005). Thus, geographic range constraint of a competent reservoir host, determined by the host’s ecological niche, is a potential driver of the constrained spatial pattern of pathogen positive ticks observed in this study. Furthermore, environmental conditions may mediate bacterial replication and transmission cycles, limiting where arthropod vectors can successfully acquire or transmit new infections (Galletti et al. 2013). Of note, while we estimated an ‘infected niche’, these ticks tested positive for the pathogen, but even with this nuanced information available, we do not precisely know if they were capable of onward transmission. In the Rocky Mountain wood tick, *Dermacentor andersoni*, there is evidence that ticks infected with *R. rickettsii* have reduced fitness (Niebylski et al. 1999). However, there are other tick-pathogen relationships that may increase fitness, such as the blacklegged tick, *Ixodes scapularis*, and the bacteria *Anaplasma phagocytophilum*, that causes the tick to express anti-freeze-like protein to enhance its survival in colder climates (Neelakanta et al. 2010). If there is an interaction between pathogens and tick fitness, this may impact the predicted suitable habitat for the infected vector.

Discrepancies in predicted geographic ranges are expected to arise as a result of the inherent differences in SDM algorithms. While there is considerable overlap in the projected output across methods in this study, low probabilities of occurrence are abundant throughout much of the projected ranges. These finding contrast with other published distributions of *D. variabilis*, where MaxEnt models have projected high bioclimatic suitability throughout the range, particularly in the central and southern United States (James et al. 2015, Minigan et al. 2018, Boorgula et al. 2020). Model results are also subject to differences in data inputs and limitations of data sampling, such as when the geographic extent of collections is subject to bias. This study was limited to data associated with a military installation reporting, and thus contained repeated observations at some locations – a common finding in surveillance geolocation data. Spatial thinning of occurrence records addresses potential oversampling of localities in these data, as many reported bites occurred in the vicinity of reporting military installations and medical treatment facilities. The thinned data used in model building is an appropriate representation of the full dataset of tick occurrences collected at clinics across the study area, where sufficient locations were represented while controlling for repeated observations (SFig. 3). Nevertheless, this does not address issues of potential under sampling across the full range of suitable habitats, or across life-stages, which could be underrepresented in this particular surveillance system. The majority of *D. variabilis* surveillance data are adult ticks collected through field sampling or found biting humans. Juvenile life stages of *D. variabilis* are rarely collected through standard field sampling methods of flagging or dragging because they live off-host, in areas that are not sampled, such as rodent burrows (Sonenshine 1993). The bias of using data from sampling methods which collect a majority of adult *D. variabilis*, may confound our understanding of predicted *D. variabilis* ranges and the dynamics of *R. montanensis*. More data are needed from all life stages to create better understanding of tick-rickettsia range distribution.

Interpolated bioclimatic variables are the primary input environmental predictors used frequently throughout the SDM literature, but they may not fully describe geographically limiting factors when modeling generalist tick species. Due to the prevalence of SDM studies that used BioClim data, we included bioclimatic layers in this study for comparison with other published models. However, few of these environmental variables independently contributed to predictions of *D. variabilis* presence, and those that were identified as underlying drivers for averaged model output were related to seasonality. When building SDMs for ticks, identification of better environmental predictors may be necessary, such as those related to soil moisture. While we included some proxies of soil conditions, they did not appreciably contribute to estimates for *D. variabilis* presence. The Normalized Difference Vegetation Index (NDVI), a quantification of surface vegetation using remote sensing measurements, has been used in other studies of tick distributions to approximate conditions that may be limiting to questing ticks. However, the importance of the contribution of NDVI to models in the literature has been mixed. We therefore suggest that future studies include further exploration of additional geospatial data layers and tick-host interactions, to better capture the environmental limits to tick and pathogen positive tick distributions, in order to use SDM approaches effectively.

This study leveraged a unique surveillance dataset to assess differences in geographic distributions between *D. variabilis* ticks, and those infected with *R. montanensis*. We have consistently demonstrated that presence of a potential vector does not inherently imply presence of the pathogen, across a range of modeling methods. This has important implications for public health agencies, which may use SDMs of vectors to infer risk and make management decisions. Moving forward, future research that uses expanded georeferenced tick surveillance data, with accompanying seroprevalence screening, will help reconcile our distribution maps with the full range of *D. variabilis* in the United States. Additional data points will allow the opportunity to better assess model performance with independent validation data that control for spatial autocorrelation, as the use of holdout data can lead to inflated model accuracy metrics (Bahn and McGill 2013). Although RF was the best performing modeling algorithm in this study, methodological choices should be made with specific goals in mind, and we caution against extrapolating the model performance shown here to other datasets or geographic foci.

## Conclusion

There is considerable overlap in the estimated geographic range of *D. variabilis* across modeling methods used in this study. Nevertheless, by conserving input data layers across modeling approaches, we demonstrated that differences in these predictions can arise as an artefact of methodology. These discrepancies in predicted range may be quite profound in impact and interpretation, depending on the intended application of results:for example, if these mapped model outcomes are used for communicating either tick encounter or disease risk at sub-regional scales. Further, we find that the predicted “infected niche” is smaller than the overall predicted tick niche, and thus predicted vector distributions may not best reflect human risk of acquiring a vector-borne disease. We therefore recommend caution in relying on single method SDMs, or those that imply disease risk from vector niches, to inform public health operations.

## Data Availability

all data are publicly available as stated in the paper itself

## Acknowledgments

A.L.R. and H.K.S. are federal/contracted employee of the United States government. This work was prepared as part of their official duties. Title 17 U.S.C. 105 provides that ’copyright protection under this title is not available for any work of the United States Government.’ Title 17 U.S.C. 101 defines a U.S. Government work as work prepared by a military service member or employee of the U.S. Government as part of that person’s official duties.

## Funding

CAL, HDG, and SJR were funded by NIH 1R01AI136035-01. ALW and SJR were additionally funded by CDC grant 1U01CK000510-01: Southeastern Regional Center of Excellence in Vector-Borne Diseases: The Gateway Program. This project was also funded by the Department of Defense Global Emerging Infections System (GEIS), work unit 000188M.0931.001.A0074. This publication was supported by the Cooperative Agreement Number above from the Centers for Disease Control and Prevention. Its contents are solely the responsibility of the authors and do not necessarily represent the official views of the Centers for Disease Control and Prevention. The views expressed in this article reflect the results of research conducted by the author and do not necessarily reflect the official policy or position of the Department of the Navy, Department of Defense, nor the United States Government.

## Supplemental Materials

**Table S1.**
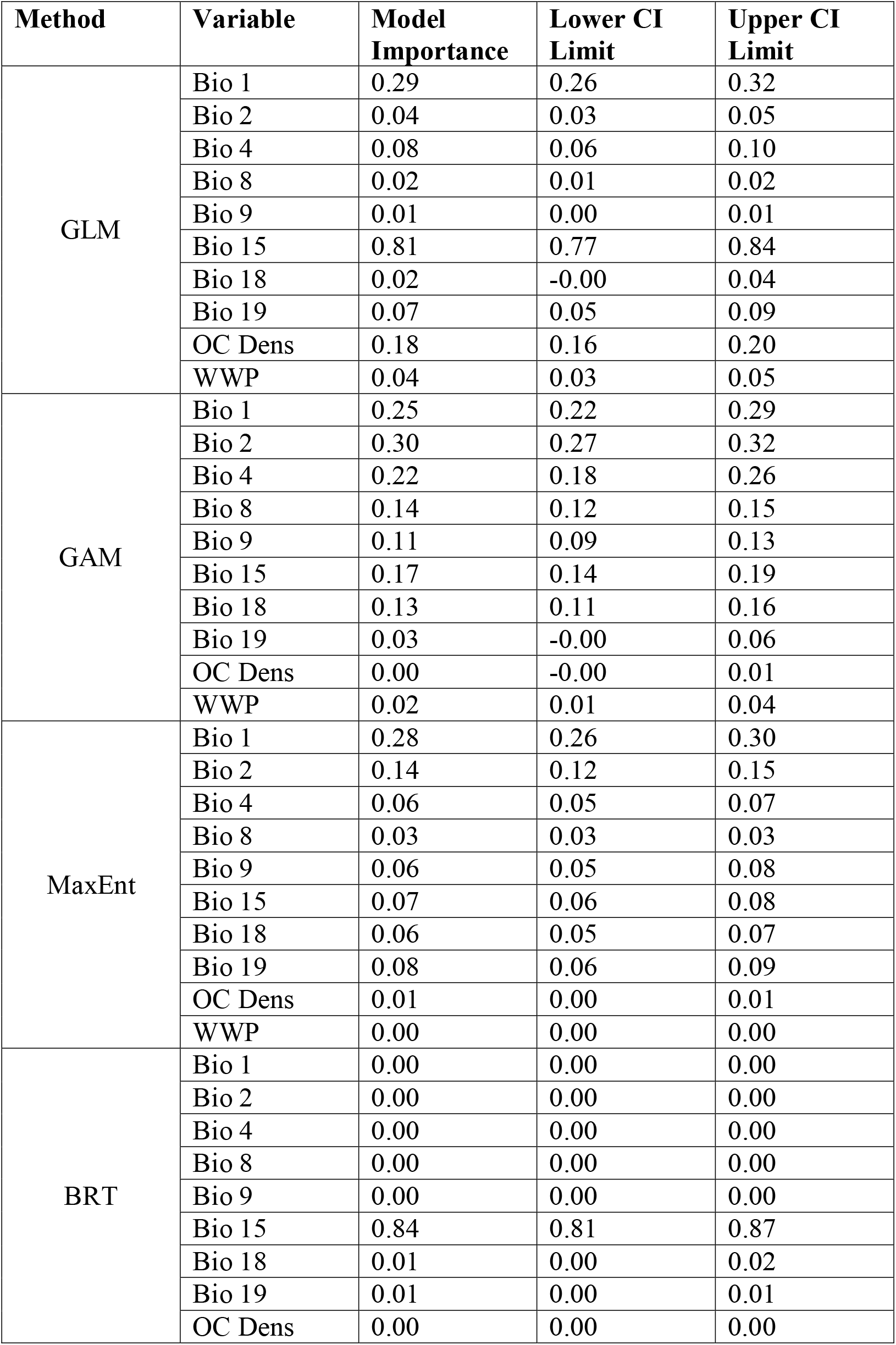
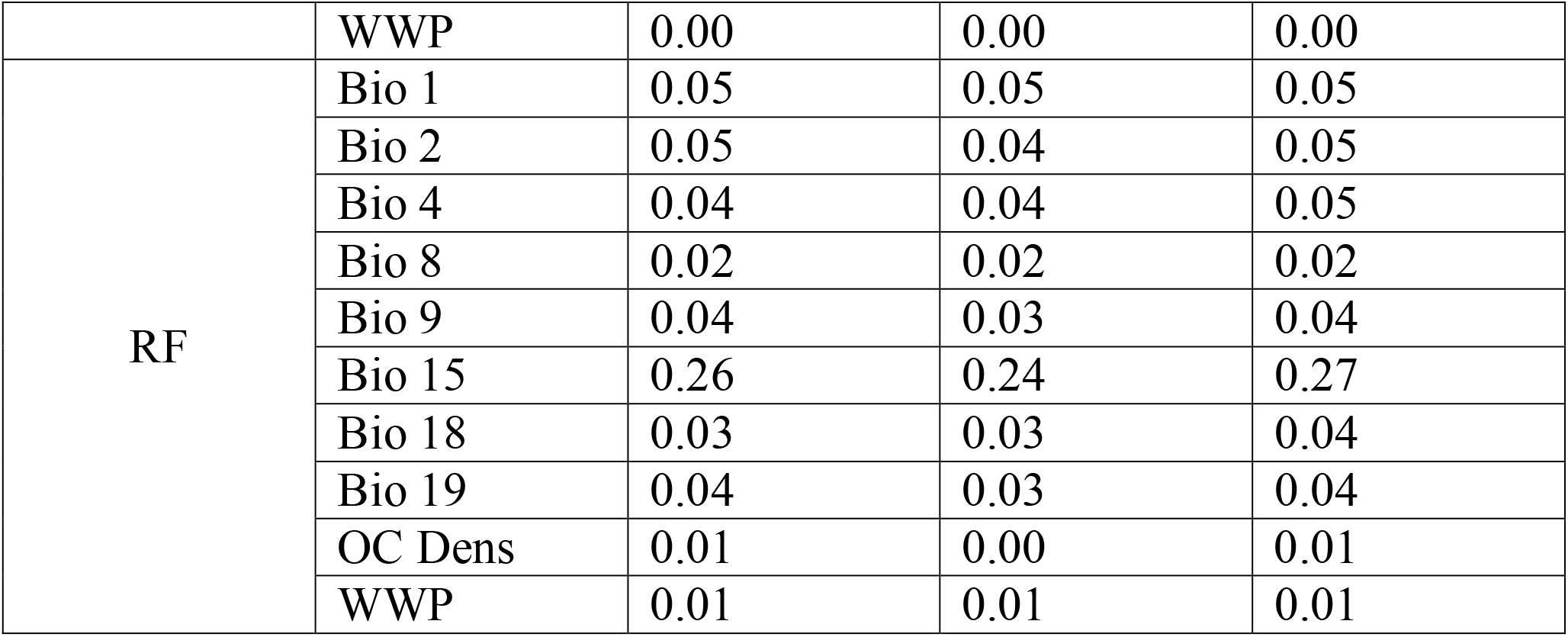
Model variable importance with confidence interval limits for models built with all *Dermacentor variabilis* ticks.

| Method | Variable | Model Importance | Lower CI Limit | Upper CI Limit |
| --- | --- | --- | --- | --- |
| GLM | Bio 1 | 0.29 | 0.26 | 0.32 |
|  | Bio 2 | 0.04 | 0.03 | 0.05 |
|  | Bio 4 | 0.08 | 0.06 | 0.10 |
|  | Bio 8 | 0.02 | 0.01 | 0.02 |
|  | Bio 9 | 0.01 | 0.00 | 0.01 |
|  | Bio 15 | 0.81 | 0.77 | 0.84 |
|  | Bio 18 | 0.02 | -0.00 | 0.04 |
|  | Bio 19 | 0.07 | 0.05 | 0.09 |
|  | OC Dens | 0.18 | 0.16 | 0.20 |
|  | WWP | 0.04 | 0.03 | 0.05 |
| GAM | Bio 1 | 0.25 | 0.22 | 0.29 |
|  | Bio 2 | 0.30 | 0.27 | 0.32 |
|  | Bio 4 | 0.22 | 0.18 | 0.26 |
|  | Bio 8 | 0.14 | 0.12 | 0.15 |
|  | Bio 9 | 0.11 | 0.09 | 0.13 |
|  | Bio 15 | 0.17 | 0.14 | 0.19 |
|  | Bio 18 | 0.13 | 0.11 | 0.16 |
|  | Bio 19 | 0.03 | -0.00 | 0.06 |
|  | OC Dens | 0.00 | -0.00 | 0.01 |
|  | WWP | 0.02 | 0.01 | 0.04 |
| MaxEnt | Bio 1 | 0.28 | 0.26 | 0.30 |
|  | Bio 2 | 0.14 | 0.12 | 0.15 |
|  | Bio 4 | 0.06 | 0.05 | 0.07 |
|  | Bio 8 | 0.03 | 0.03 | 0.03 |
|  | Bio 9 | 0.06 | 0.05 | 0.08 |
|  | Bio 15 | 0.07 | 0.06 | 0.08 |
|  | Bio 18 | 0.06 | 0.05 | 0.07 |
|  | Bio 19 | 0.08 | 0.06 | 0.09 |
|  | OC Dens | 0.01 | 0.00 | 0.01 |
|  | WWP | 0.00 | 0.00 | 0.00 |
| BRT | Bio 1 | 0.00 | 0.00 | 0.00 |
|  | Bio 2 | 0.00 | 0.00 | 0.00 |
|  | Bio 4 | 0.00 | 0.00 | 0.00 |
|  | Bio 8 | 0.00 | 0.00 | 0.00 |
|  | Bio 9 | 0.00 | 0.00 | 0.00 |
|  | Bio 15 | 0.84 | 0.81 | 0.87 |
|  | Bio 18 | 0.01 | 0.00 | 0.02 |
|  | Bio 19 | 0.01 | 0.00 | 0.01 |
|  | OC Dens | 0.00 | 0.00 | 0.00 |
|  | WWP | 0.00 | 0.00 | 0.00 |
| RF | Bio 1 | 0.05 | 0.05 | 0.05 |
|  | Bio 2 | 0.05 | 0.04 | 0.05 |
|  | Bio 4 | 0.04 | 0.04 | 0.05 |
|  | Bio 8 | 0.02 | 0.02 | 0.02 |
|  | Bio 9 | 0.04 | 0.03 | 0.04 |
|  | Bio 15 | 0.26 | 0.24 | 0.27 |
|  | Bio 18 | 0.03 | 0.03 | 0.04 |
|  | Bio 19 | 0.04 | 0.03 | 0.04 |
|  | OC Dens | 0.01 | 0.00 | 0.01 |
|  | WWP | 0.01 | 0.01 | 0.01 |

**Table S2.** Model variable importance with confidence interval limits for models built with the subset of ticks infected with *R. montanensis*.

| Method | Variable | Model Importance | Lower CI Limit | Upper CI Limit |
| --- | --- | --- | --- | --- |
| GLM | Bio 1 | 0.63 | 0.60 | 0.66 |
|  | Bio 2 | 0.24 | 0.20 | 0.27 |
|  | Bio 4 | 0.60 | 0.57 | 0.62 |
|  | Bio 8 | 0.07 | 0.04 | 0.10 |
|  | Bio 9 | 0.15 | 0.10 | 0.20 |
|  | Bio 15 | 0.99 | 0.98 | 0.99 |
|  | Bio 18 | 0.51 | 0.46 | 0.56 |
|  | Bio 19 | 0.40 | 0.35 | 0.44 |
|  | OC Dens | 0.05 | 0.02 | 0.07 |
|  | WWP | 0.26 | 0.21 | 0.31 |
| GAM | Bio 1 | 0.41 | 0.25 | 0.58 |
|  | Bio 2 | 0.28 | 0.20 | 0.36 |
|  | Bio 4 | 0.46 | 0.35 | 0.57 |
|  | Bio 8 | 0.13 | 0.02 | 0.24 |
|  | Bio 9 | 0.22 | 0.06 | 0.38 |
|  | Bio 15 | 0.68 | 0.58 | 0.77 |
|  | Bio 18 | 0.50 | 0.39 | 0.61 |
|  | Bio 19 | 0.26 | 0.13 | 0.38 |
|  | OC Dens | 0.10 | 0.01 | 0.18 |
|  | WWP | 0.17 | 0.07 | 0.26 |
| MaxEnt | Bio 1 | 0.37 | 0.31 | 0.43 |
|  | Bio 2 | 0.18 | 0.15 | 0.21 |
|  | Bio 4 | 0.33 | 0.29 | 0.37 |
|  | Bio 8 | 0.02 | 0.00 | 0.03 |
|  | Bio 9 | 0.01 | 0.00 | 0.02 |
|  | Bio 15 | 0.72 | 0.69 | 0.75 |
|  | Bio 18 | 0.27 | 0.22 | 0.32 |
|  | Bio 19 | 0.09 | 0.06 | 0.11 |
|  | OC Dens | 0.01 | 0.00 | 0.01 |
|  | WWP | 0.07 | 0.05 | 0.09 |
| BRT | Bio 1 | 0.00 | 0.00 | 0.00 |
|  | Bio 2 | 0.00 | 0.00 | 0.00 |
|  | Bio 4 | 0.00 | 0.00 | 0.00 |
|  | Bio 8 | 0.00 | 0.00 | 0.00 |
|  | Bio 9 | 0.00 | 0.00 | 0.00 |
|  | Bio 15 | 0.98 | 0.97 | 0.99 |
|  | Bio 18 | 0.00 | 0.00 | 0.00 |
|  | Bio 19 | 0.00 | 0.00 | 0.00 |
|  | OC Dens | 0.00 | 0.00 | 0.00 |
|  | WWP | 0.00 | 0.00 | 0.00 |
| RF | Bio 1 | 0.04 | 0.03 | 0.04 |
|  | Bio 2 | 0.05 | 0.04 | 0.05 |
|  | Bio 4 | 0.05 | 0.04 | 0.05 |
|  | Bio 8 | 0.04 | 0.04 | 0.05 |
|  | Bio 9 | 0.02 | 0.02 | 0.02 |
|  | Bio 15 | 0.25 | 0.23 | 0.27 |
|  | Bio 18 | 0.04 | 0.03 | 0.04 |
|  | Bio 19 | 0.04 | 0.04 | 0.05 |
|  | OC Dens | 0.01 | 0.00 | 0.01 |
|  | WWP | 0.03 | 0.02 | 0.04 |

**Fig. S1.**
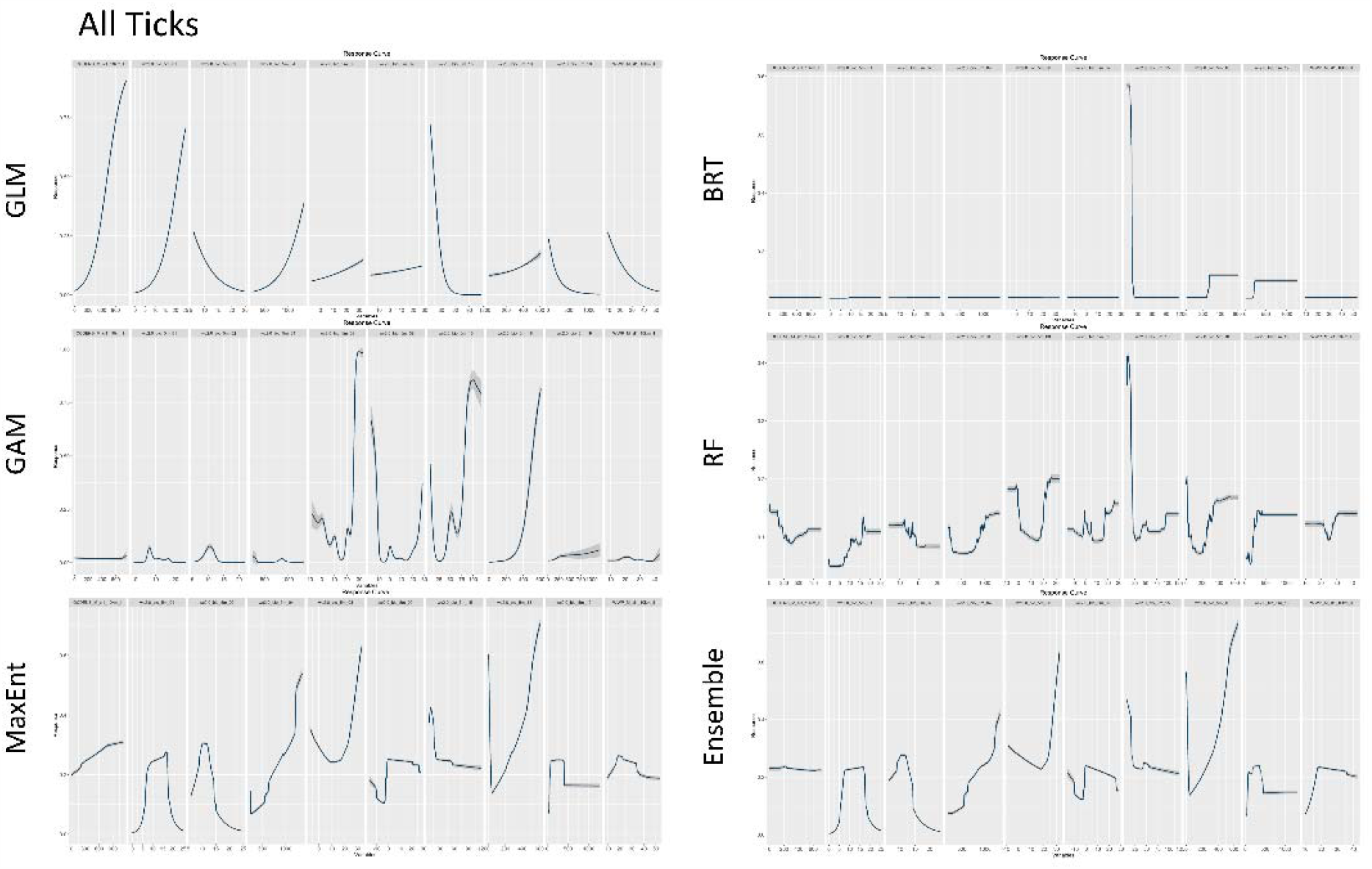
Averaged variable response curves, with 95% confidence intervals, for environmental variables used to build species distribution models (SDMs) with the full dataset of *D. variabilis* occurrences. The variables that influenced the predicted presence of ticks varied between the modeling methods used, including generalized linear model (GLM), generalized additive model (GAM), maximum entropy (MaxEnt), boosted regression trees (BRT), and random forests (RF), and a weighted ensemble of these five methods.

**Fig. S2.**
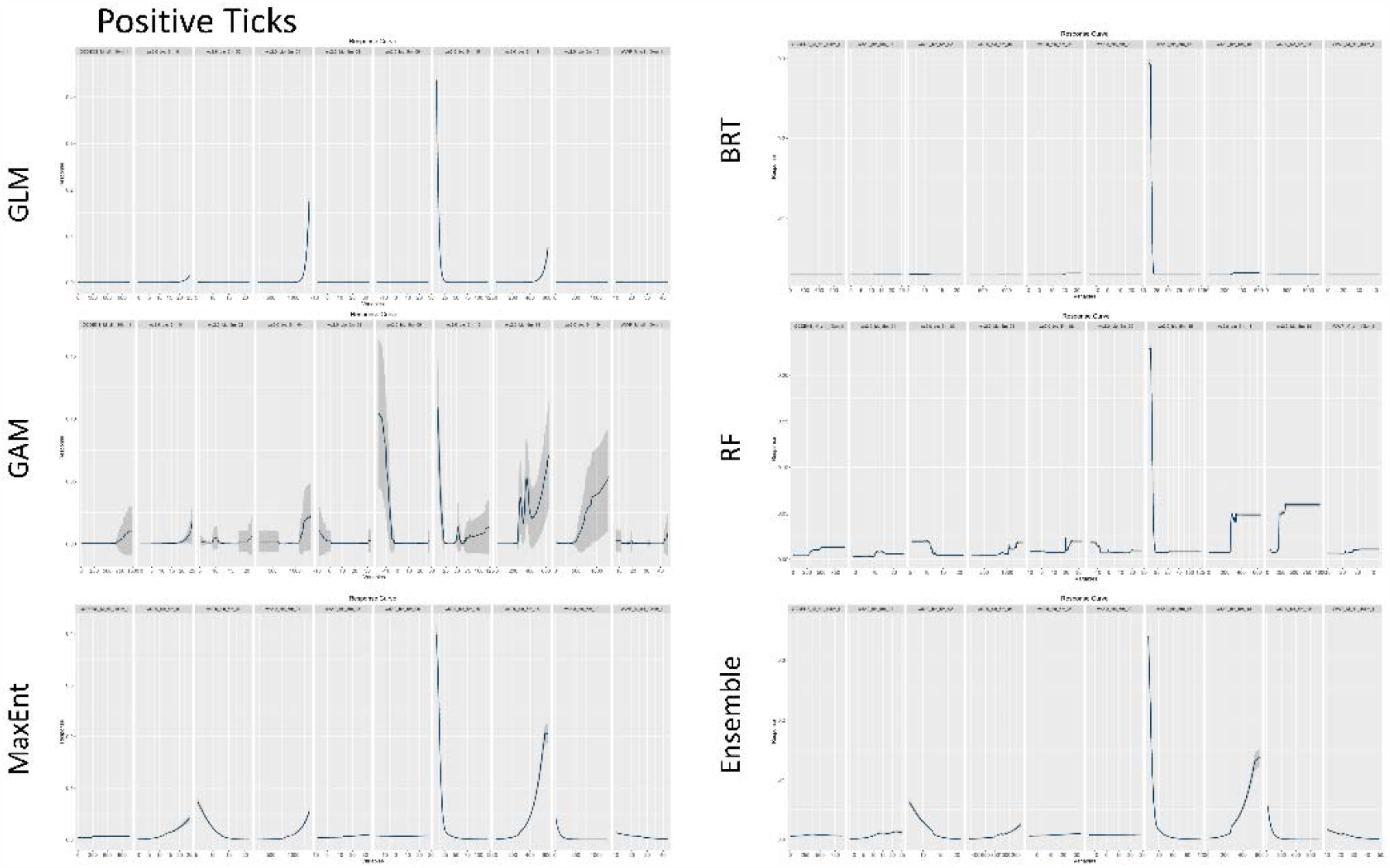
Averaged variable response curves, with 95% confidence intervals, for environmental variables used to build species distribution models (SDMs) with the subset of *D. variabilis* ticks infected with *R. montanensis*. The variables that influenced the predicted presence of infected ticks varied between the modeling methods used, including generalized linear model (GLM), generalized additive model (GAM), maximum entropy (MaxEnt), boosted regression trees (BRT), and random forests (RF), and a weighted ensemble of these five methods.

**Fig. S3.**
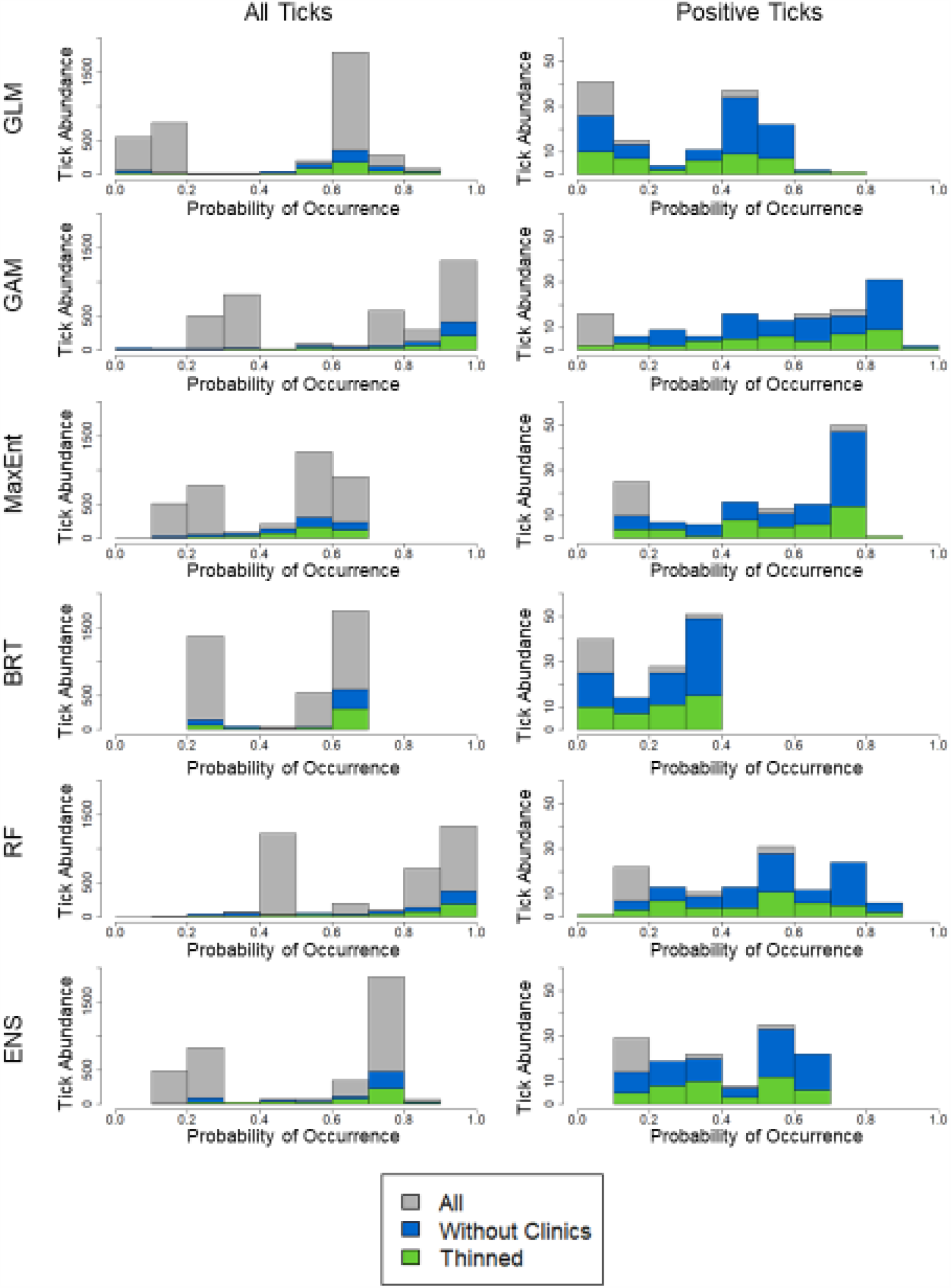
Abundance of tick occurrence points shown against predicted probability of occurrence across modeling methods for the full dataset (grey), data with repeated observations at reporting clinic locations removed (blue), and spatially thinned data used in SDM building (green). All ticks include all *Dermacentor variabilis* ticks, and positive ticks are *D. variabilis* ticks that tested positive for *Rickettsia montanensis* infections.

## Notes

### Competing Interest Statement

The authors have declared no competing interest.

## References

Aiello-Lammens, M. E., R. A. Boria, A. Radosavljevic, B. Vilela, and R. P. Anderson. 2015. spThin: an R package for spatial thinning of species occurrence records for use in ecological niche models. Ecography. 38: 541–545.

Araujo, M., and M. New. 2007. Ensemble forecasting of species distributions. Trends in Ecology & Evolution. 22: 42–47.

Bahn, V., and B. J. McGill. 2013. Testing the predictive performance of distribution models. Oikos. 122: 321–331.

Baldridge, G. D., N. Y. Burkhardt, A. S. Oliva, T. J. Kurtti, and U. G. Munderloh. 2010. Rickettsial ompB promoter regulated expression of GFPuv in transformed Rickettsia montanensis. PloS one. 5: e8965.

Blackburn, J. K., S. Matakarimov, S. Kozhokeeva, Z. Tagaeva, L. K. Bell, I. T. Kracalik, and A. Zhunushov. 2017. Modeling the ecological niche of Bacillus anthracis to map anthrax risk in Kyrgyzstan. Am. J. Trop. Med. Hyg. 96: 550–556.

Boorgula, G. D. Y., A. T. Peterson, D. H. Foley, R. R. Ganta, and R. K. Raghavan. 2020. Assessing the current and future potential geographic distribution of the American dog tick, Dermacentor variabilis (Say) (Acari: Ixodidae) in North America. PLOS ONE. 15: e0237191.

Breiman, L. 2001. Random Forests. Machine Learning. 45: 5–32.

Burtis, J. C., J. B. Yavitt, T. J. Fahey, and R. S. Ostfeld. 2019. Ticks as soil-dwelling arthropods: an intersection between disease and soil ecology. Journal of Medical Entomology. 56: 1555–1564.

Carlson, C. J., E. Dougherty, M. Boots, W. Getz, and S. J. Ryan. 2018. Consensus and conflict among ecological forecasts of Zika virus outbreaks in the United States. Scientific Reports. 8.

CDC. 2010. Spotted Fever Rickettsiosis (Rickettsia spp.) 2010 Case Definition (No. CSTE Position Statement: 09-ID-16). Centers for Disease Control and Prevention.

CDC. 2019a. Rocky Mountain Spotted Fever (RMSF) Epidemiology and Statistics.

CDC. 2019b. Regions where ticks live.

Chatterjee, S., and A. S. Hadi. 2006. Analysis of Collinear Data, pp. 221–258. In Regression Analysis by Example. John Wiley & Sons, Inc., Hoboken, NJ, USA.

De Marco, P., and C.C. Nóbrega. 2018. Evaluating collinearity effects on species distribution models: An approach based on virtual species simulation. PLOS ONE. 13: e0202403.

Elith, J., and J. R. Leathwick. 2009. Species Distribution Models: Ecological Explanation and Prediction Across Space and Time. Annual Review of Ecology, Evolution, and Systematics. 40: 677–697.

Elith, J., J. R. Leathwick, and T. Hastie. 2008. A working guide to boosted regression trees. Journal of Animal Ecology. 77: 802–813.

ESRI. 2016. ArcGIS 10.4. Environmental Systems Research Institute (ESRI), Redlands, CA.

Evans, J. S., M. A. Murphy, Z. A. Holden, and S. A. Cushman. 2011. Modeling Species Distribution and Change Using Random Forest, pp. 139–159. In Drew, C.A., Wiersma, Y.F., Huettmann, F. (eds.), Predictive Species and Habitat Modeling in Landscape Ecology. Springer New York, New York, NY.

Fick, S. E., and R. J. Hijmans. 2017. WorldClim 2: new 1_Jkm spatial resolution climate surfaces for global land areas. International Journal of Climatology. 37: 4302–4315.

Galletti, M. F. B. M., A. Fujita, M Y. Nishiyama Jr, C. D. Malossi, A. Pinter, J. F. Soares, S. Daffre, M. B. Labruna, and A.C. Fogaça. 2013. Natural Blood Feeding and Temperature Shift Modulate the Global Transcriptional Profile of Rickettsia rickettsii Infecting Its Tick Vector. PLoS ONE. 8: e77388.

GDAL/OGR contributors. 2020. GDAL/OGR Geospatial Data Abstraction software Library. Open Source Geospatial Foundation.

Gurgel-Gonçalves, R., C. Galvão, J. Costa, and A. T. Peterson. 2012. Geographic distribution of Chagas disease vectors in Brazil based on ecological niche modeling. Journal of Tropical Medicine. 2012: 1–15.

Hao, T., J. Elith, G. Guillera□Arroita, and J.J. Lahoz□Monfort. 2019. A review of evidence about use and performance of species distribution modelling ensembles like BIOMOD. Diversity and Distributions. 25: 839–852.

Hardstone Yoshimizu, M., and S. A. Billeter. 2018. Suspected and confirmed vector-borne rickettsioses of North America associated with human diseases. Tropical Medicine and Infectious Disease. 3: 2.

Hengl, T., J. Mendes de Jesus, G. B. M. Heuvelink, M. Ruiperez Gonzalez, M. Kilibarda, A. Blagotić, W. Shangguan, M. N. Wright, X. Geng, B. Bauer-Marschallinger, M. A. Guevara, R. Vargas, R. A. MacMillan, N. H. Batjes, J. G. B. Leenaars, E. Ribeiro, I. Wheeler, S. Mantel, and B. Kempen. 2017. SoilGrids250m: Global gridded soil information based on machine learning. PLOS ONE. 12: e0169748.

James, A. M., C. Burdett, M. J. Mccool, A. Fox, and P. Riggs. 2015. The geographic distribution and ecological preferences of the American dog tick, Dermacentor variabilis (Say), in the U.S.A. Medical and Veterinary Entomology. 29: 178–188.

Lippi, C. A., A. M. Stewart-Ibarra, M. E. F. B. Loor, J. E. D. Zambrano, N. A. E. Lopez, J.K. Blackburn, and S. J. Ryan. 2019. Geographic shifts in Aedes aegypti habitat suitability in Ecuador using larval surveillance data and ecological niche modeling: Implications of climate change for public health vector control. PLOS Neglected Tropical Diseases. 13: e0007322.

McCullagh, P., and J. A. Nelder. 1998. Generalized linear models, 2nd ed. ed, Monographs on statistics and applied probability. Chapman & Hall/CRC, Boca Raton.

McQuiston, J. H., G. Zemtsova, J. Perniciaro, M. Hutson, J. Singleton, W. L. Nicholson, and M. L. Levin. 2012. Afebrile spotted fever group Rickettsia infection after a bite from a Dermacentor variabilis tick infected with Rickettsia montanensis. Vector-Borne and Zoonotic Diseases. 12: 1059–1061.

Merow, C., M. J. Smith, and J. A. Silander. 2013. A practical guide to MaxEnt for modeling species’ distributions: what it does, and why inputs and settings matter. Ecography. 36: 1058–1069.

Minigan, J. N., H. A. Hager, A. S. Peregrine, and J. A. Newman. 2018. Current and potential future distribution of the American dog tick (Dermacentor variabilis, Say) in North America. Ticks and Tick-borne Diseases. 9: 354–362.

Morales, N. S., I. C. Fernández, and V. Baca-González. 2017. MaxEnt’s parameter configuration and small samples: are we paying attention to recommendations? A systematic review. PeerJ. 5: e3093.

Naimi, B., and M.B. Araújo. 2016. sdm: a reproducible and extensible R platform for species distribution modelling. Ecography. 39: 368–375.

Neelakanta, G., H. Sultana, D. Fish, J. F. Anderson, and E. Fikrig. 2010. Anaplasma phagocytophilum induces Ixodes scapularis ticks to express an antifreeze glycoprotein gene that enhances their survival in the cold. The Journal of Clinical Investigation. 120: 3179–3190.

Nicholson, W. L., and C. D. Paddock. 2019. Rickettsial Diseases (Including Spotted Fever & Typhus Fever Rickettsioses, Scrub Typhus, Anaplasmosis, and Ehrlichioses). In CDC Yellow Book. Centers for Disease Control and Prevention.

Niebylski, M. L., M. G. Peacock, and T. G. Schwan. 1999. Lethal effect of Rickettsia rickettsii on its tick vector (Dermacentor andersoni). Appl. Environ. Microbiol. 65: 773–778.

Parola, P., C. D. Paddock, and D. Raoult. 2005. Tick-borne rickettsioses around the world: emerging diseases challenging old concepts. Clinical Microbiology Reviews. 18: 719– 756.

Peterson, A. T., and J. Soberón. 2012. Species Distribution Modeling and Ecological Niche Modeling: Getting the Concepts Right. Natureza & Conservação. 10: 102–107.

Phillips, S. J., and M. Dudík. 2008. Modeling of species distributions with Maxent: new extensions and a comprehensive evaluation. Ecography. 31: 161–175.

R Core Team. 2019. R: A language and environment for statistical computing. R Foundation for Statistical Computing, Vienna, Austria.

Sonenshine, D. E. 1993. Biology of Ticks. Oxford University Press, New York.

St. John, H.K., M. L. Adams, P. M. Masuoka, J. G. Flyer-Adams, J. Jiang, P. J. Rozmajzl, E. Y. Stromdahl, and A. L. Richards. 2016. Prevalence, distribution, and development of an ecological niche model of Dermacentor variabilis ticks positive for Rickettsia montanensis. Vector-Borne and Zoonotic Diseases. 16: 253–263.

Townsend Peterson, A., M. Papeş, and M. Eaton. 2007. Transferability and model evaluation in ecological niche modeling: a comparison of GARP and Maxent. Ecography. 30: 550– 560.

Wood, S. N. 2006. Generalized additive models: an introduction with R, Texts in statistical science. Chapman & Hall/CRC, Boca Raton, FL.

